# Adherence to prophylaxis and bleeding outcome: a multicenter Nigerian study

**DOI:** 10.1101/2022.02.14.22270964

**Authors:** Theresa U. Nwagha, Helen C. Okoye, Saleh Yuguda, Christiana E. Udo, Mutiat K Ogunfemi, Dalhat H. Gwarzo, Joel N. Osuji

## Abstract

**Background:** In Nigeria, low-dose prophylaxis is the standard of care as it reduces bleeding, development of target joints, arthropathy, and improvement of quality of life. Non-adherence or poor adherence can prevent the achievement of these outcomes. The levels and determinants of (non-)adherence among persons with haaemophilia (PWH) in Sub-Saharan Africa have not been evidenced.

**Objective:** To evaluate self-reported adherence among PWH, provide evidence of determinants/predictors of adherence and establish the associations between nonadherence and presence of target joints and annualized bleed rate.

**Methodology:** A cross-sectional survey of 42 participants on low-dose prophylaxis recruited during outpatient appointments in 5 haemophilia treatment centers in Nigeria. We used the validated Haemophilia Regimen Treatment Adherence Scale-Prophylaxis (VERITAS -Pro), 24 questions on six subscales (time, dose, plan, remember, skip, and communicate) questionnaire. The options of VERITAS -Pro were represented in a 5 Likert scale and the possible subscale ranged from 4 points (most adherent) to 20 points (least adherent) and the possible total score ranged from 24 (most adherent) to 120 (least adherent) the cutoff for overall adherence put at > 61 to indicate nonadherence. Information on the presence of target joints, the number of target joints, and annualized bleeding rates were collected from medical files.

**Results:** The mean age of the participants was 9.79 (6.29) years, with 96.6% having hemophilia A and 79.3% having target joints. Overall adherence to the prophylaxis regimen was 81.0%. The mean total VERITAS-Pro for the adherent group and the non-adherent group was 37.35 ±9.08 and 63.0± 6.37, respectively. The mean subscale scores for the adherent group ranged from 0.67 (communication) to 8.68 (planning), while the mean subscale scores range from 1.0 communication to 13.88 (planning) for the nonadherent group. The mean difference of all except the dosing subscale was statistically significant with p<0.05. Only the skipping subscale showed a statistically significant positive correlation with ABR in the non-adherent group p=0.02.

**Conclusions:** The findings indicate that adherence was very good, and most were in communication with their treatment centers. The skipping subscale was significantly associated with ABR for the nonadherent group. Interventions aimed at improving adherence are the key to better treatment outcomes. A multicenter study was needed to assess the reason for poor adherence.

## Introduction

Prophylaxis is defined as treatment by regular routine intravenous infusions of factor concentrate with the goal of preventing anticipated bleeding episodes.^1^ It is the standard of care in hemophilia. Prophylaxis was conceived from the observation that moderate hemophilia patients with clotting factor level >1 IU/dl seldom experience spontaneous bleeding and have much better preservation of joint function ^1^ Prophylactic replacement of clotting factor has been shown to be useful even when factor levels are not maintained above 1 IU/dl at all-time ^3^

Prophylaxis in children is most efficient when started at an early age.^4^ The key to a successful long-term outcome in patients with hemophilia is an efficient prophylaxis that prevents bleeding in joints for children and adults with hemophilia. Prophylaxis does not reverse established joint damage; however, it decreases frequency of bleeding and may slow progression of joint disease and improve quality of life. Efficient prophylaxis requires taking into account the available resources which includes the clotting factor concentrate, trough levels, the bleeding trigger (activity levels, chronic synovitis, already existing arthropathy), and most importantly the number of acceptable bleeds, especially joint bleeds.^5^

Depending on the available resources, the treatment objectives can vary between countries and treatment centres. In countries with significant resource constraints, lower doses of prophylaxis given more frequently may be an effective option. In an almost ideal setting, the number of spontaneous bleeds should be minimized to prevent the manifestation of joint arthropathy. Prophylaxis has greatly improved joint health and its challenging joint outcome assessment.^5,6^

There are three types of prophylaxis primary, secondary and tertiary. Primary prophylaxis is a regular continuous prophylaxis started in the absence of documented joint disease, determined by physical examination and/or imaging studies, and before the second clinically evident joint bleed and 3 years of age. The use of primary prophylaxis has allowed many children with severe hemophilia to live more normal lives with fewer acute bleeding episodes and decreased orthopedic complications When begun at an early age and continued in an uninterrupted fashion. primary prophylaxis has been useful in preventing hemophilic arthropathy. Secondary prophylaxis is regular continuous prophylaxis initiated after 2 or more joint bleeds but before the onset of joint disease; this is usually at 3 or more years of age. Tertiary prophylaxis is regular continuous prophylaxis initiated after the onset of documented joint disease. Tertiary prophylaxis typically applies to prophylaxis commenced in adulthood.^7^

How to start and when to start prophylaxis with either standard half-life (SHL) or extended half-life (EHL) Coagulation factor concentrates (CFCs) are not significantly different. In both cases, prophylaxis should be commenced early by starting with a high-dose/high-frequency approach or a low-frequency approach, followed by escalation of frequency.^1,7^With EHL CFCs, less frequent infusions (once weekly) may be sufficient for many individuals, particularly those with severe hemophilia B receiving EHL FIX CFCs. As EHL CFCs must still be given intravenously, they remain difficult to administer in very young children with poor peripheral venous access.^8^

Depending on the patient’s age and underlying conditions, prophylaxis and its subsequent prevention of bleeds have different objectives ^9^ Prophylaxis provides protection from other types of hemorrhages in hemophilia, including preventing or substantially reducing the risk of intracranial hemorrhage.^10^ Longer-term benefits include reduction of chronic musculoskeletal pain, functional limitations and disability, need for orthopedic surgery, hospitalization, emergency room visits, and reduced length of hospital stays; all of this leads to greater participation in educational, recreational, and professional activities, with improved quality of life.^11^

In resource-constrained countries, low-dose prophylaxis tends to focus on the use of smaller doses. This is a way for patients in these countries to start receiving prophylaxis but at lower cost. To minimize cost, the focus tends to be on minimizing the doses used while keeping infusion frequencies similar.^12^ In Nigeria low dose prophylaxis is the standard of care for haemophilia as it reduces bleed, development of target joint, haemophilic arthropathy, at the same time improve quality of life.

Despite the benefits of prophylaxis, adherence has traditionally been a significant problem. There are many reasons for reduced adherence to prophylaxis. The main reason is likely the burden of administering CFCs both intravenously and frequently. This results in venous access difficulties (especially in young children but also in older adults with significant arthropathy and potentially extinguished veins) and child/family resistance to the time-consuming nature of conventional prophylaxis.

Non-adherence or poor adherence can prevent the achievement of these outcomes. Adherence among person with haemophilia on prophylaxis in Sub-Sahara Africa has not been evidenced.

## Materials and Methods

This was a cross-sectional multicenter study carried out in five (5) Haemophilia treatment centers (HTCs) in Nigeria over a period of 4 months. The participating institutions include University of Nigeria Teaching Hospital Enugu, University of Ilorin Teaching Hospital Ilorin, Aminu Kano Teaching Hospital Kano, National Hospital Abuja and Federal medical center Gombe Adherence to prophylaxis was measured using a validated Haemophilia Regimen Treatment Adherence Scale-Prophylaxis (VERITAS -Pro), 24 questions on six (four-item) subscales (time, dose, plan, remember, skip, and communicate) questionnaire which was administered to patients or their caregivers. Ethical approval for the study was gotten from the Research Ethics committee of the University of Nigeria Teaching Hospital.

### Participants

A total of 42 patients on low dose prophylaxis of clotting factor concentrates (CFC) were recruited for this study using a convenience sampling method during their outpatient visits. Inclusion criteria were all patients with haemophilia A or B on prophylaxis with CFC (self-infused or given by the caregiver) in the last one year. Patients on “on-demand” therapy regimen and those not able to infuse at home were excluded from the study. Permission to access infusion log data was sought from individuals for the purpose of this study. Participants were assured that all study-related data will be kept confidential by the study coordinator and identification will only be through an assigned unique number.

### Adherence measurement

The Validated Haemophilia Regimen Treatment Adherence Scale-Prophylaxis- (VERITAS-Pro) is a 24 -item questionnaire divided into six (6) sub scales: Time, Dose, Plan, Remember, Skip and Communicate. Response options are presented as five-point Likert scales ranging from ‘Always’ to ‘Never’; Always reflects best possible adherence for some items and the worst possible adherence for other items. Total scores range from 24-120, and subscale scores range from 4-20 with lower scores indicating higher adherence. The patient or caregiver was given the VERITAS-Pro and allowed as much time as necessary to complete the survey. Participants not returning the survey within a week received reminders via text messages or phone calls and were considered lost to follow-up after two unanswered phone calls/messages.

### Data analysis

Data collected were analyzed using the Statistical Package for Social Science (SPSS) software. Mean and standard deviations (descriptive statistics) were calculated for each subscale. Frequency and proportion were used to describe the categorical variables. Pearson correlation was used to analyze the association between self-reported adherence and annualized bleeding rate.

## Results

A total of 42 persons with haemophilia (PWH) on prophylaxis regime from 5 Haemophilia treatment centers participated in the study. The majority (96.6%) of the patients had haemophilia A. The age of the participants varied from 1 to 30 years (mean: 9.8 ± 6.3years). Thirty-three patients (79.3%) had target joints.

### Self-Reported Adherence

Adherence was defined as the total sum of all subscales <61 and nonadherence as the total sum of all subscales >61 based on the cutoff scores proposed by the original validation study^13^. There was a record of 81.0% adherence among the participants. For the subscales, 83.3% of the participants were more adherent in the timing of their prophylaxis and communication with the HTC, while the highest non-adherence,52.4% was recorded in the planning subscale (see Table 1)

**Table 1:** Adherence Status in Subscales.

| Variable | Frequency |  |
| --- | --- | --- |
| Adherence Status | (n = 42) | Percent |
| Adherent | 34 | 81.0 |
| Non-adherent | 8 | 19.0 |
| Subscales | Adherent | Non-adherent |
| Timing | 35 (83.3) | 7 (16.7) |
| Dosing | 31 (73.8) | 11 (26.2) |
| Planning | 20 (47.6) | 22 (52.4) |
| Remember | 39 (92.9) | 3 (7.1) |
| Skipping | 34 (81.0) | 8 (19.0) |
| Communication | 35 (83.3) | 7 (16.7) |

The mean logarithmic age for the adherent group was 9.77±1.62 while for the non-adherent group, it was 4.90± 2.34 this was statistically significant with a p-value of 0.02. The difference between the mean ABR and the mean number of target joints between the adherent and non-adherent groups was not statistically significant with p values of 0.90 and 0.31, respectively. (See table 2)

**Table 2:** Age, ABR and number of target joints in the adherent vs. non-adherent group.

| Variable | Adhere |  |  |  | Non-Adherence |  |  |  | t (p-value) | M. Diff |
| --- | --- | --- | --- | --- | --- | --- | --- | --- | --- | --- |
|  | Mean | SD | Min | Max | Mean | SD | Min | Max |  |  |
| Age (Log) | 9.77 | 1.62 | 3.98 | 30.19 | 4.90 | 2.34 | 1.99 | 16.98 | 2.57<br>(0.016) | 1.95 |
| Annualized Bleeding Rate (Log) | 8.12 | 2.95 | 1.99 | 60.25 | 7.59 | 6.17 | 1.99 | 60.26 | 0.13<br>(0.894) | 1.09 |
| Number of Target joint (Log) | 2.57 | 1.35 | 1.99 | 3.98 | 2.00 | 1.00 | 1.99 | 1.99 | 1.08<br>(0.308) | 1.29 |

### Overall Veritas Pro scores

The mean total VERITAS-Pro score for the adherence group for the total sample was 37.35 ± 9.08 with a range of 24 to 56 (Table 3). Subscale mean scores ranged from 0.67 (communication) to 8.6 (planning), indicating that participants reported the highest adherence to communicating with HTC and the lowest adherence to planning for administering prophylaxis. For the non-adherent group, Subscale mean scores ranged from 0.60 (communicating) to 10 (skipping), indicating that participants reported the highest adherence to communicating with HTC and the lowest adherence in skipping prophylaxis.

**Table 3:** Overall self-reported adhesion

| Variable | Adherence |  |  |  |  |  |  | Non-Adherence |  |  |  |  |  |  | t (p-value) | M. Diff |
| --- | --- | --- | --- | --- | --- | --- | --- | --- | --- | --- | --- | --- | --- | --- | --- | --- |
|  | N | Mean | SD | Min | Max | Skew | Kurt | N | Mean | SD | Min | Max | Skew | Kurt |  |  |
| Self-Reported Adherence |  |  |  |  |  |  |  |  |  |  |  |  |  |  |  |  |
| Timing | 34 | 0.78 | 0.17 | 0.60 | 1.23 | 0.67 | -0.36 | 8 | 1.02 | 0.13 | 0.85 | 1.15 | -0.16 | -2.33 | -4.43 (0.001*) | -0.24 |
| Dosing | 34 | 5.12 | 1.68 | 4 | 9 | 1.30 | 0.22 | 8 | 6.25 | 2.49 | 4 | 10 | 0.29 | -1.98 | -1.56 (0.127) | -1.13 |
| Planning | 34 | 8.68 | 3.23 | 4 | 17 | 0.68 | 0.43 | 8 | 13.88 | 4.49 | 5 | 20 | -1.08 | 1.76 | -3.10 (0.013*) | -5.19 |
| Remembering | 34 | 5.62 | 1.94 | 4 | 10 | 0.95 | -0.48 | 8 | 8.25 | 2.44 | 6 | 13 | 1.04 | 0.91 | -2.85 (0.019*) | -2.63 |
| Skipping | 34 | 6.50 | 2.53 | 4 | 13 | 0.73 | -0.38 | 8 | 12.88 | 2.64 | 10 | 17 | 0.49 | -1.13 | -6.19 (<0.001*) | -6.38 |
| Communicating | 34 | 0.67 | 0.13 | 0.60 | 1.15 | 2.16 | 4.82 | 8 | 1.00 | 0.18 | 0.60 | 1.15 | -1.85 | 3.54 | -4.91 (0.001*) | -0.34 |
| Total | 34 | 37.35 | 9.08 | 24 | 56 | 0.46 | -0.61 | 8 | 63.00 | 6.37 | 58 | 78 | 2.30 | 5.88 | 14.59 (<0.001*) | -25.65 |

SD = standard deviation \* statistical significance

There was a significant difference in the overall VERITAS-Pro score between the adherence group and the non-adherent group, p<0.01. For the subscale, there was a significant difference in timing, planning communication, skipping, and remembering with p< 0.05 See Table 3.

### Correlation analysis of self-reported adherence, annualized bleeding rate, and number of target joint

The association between self-reported adherence and annualized bleeding rate was tested with Pearson’s correlation, which revealed that there were non-statistically significant associations with subscales between the group except for ‘Skipping’ among the non-adherent group, with a significantly high positive correlation coefficient of 0.94 and a p-value of 0.018; See Table 4.

**Table 4:**
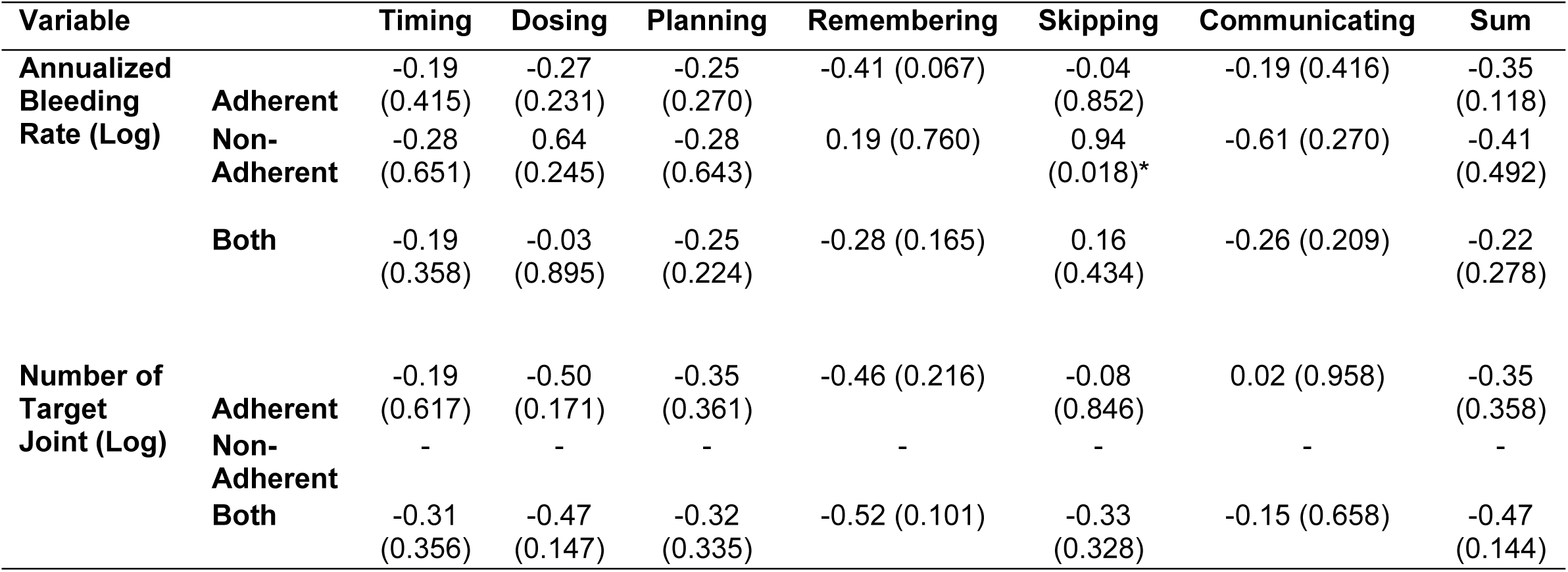
Association between self-reported adherence, annualized bleeding rate, and Number of Target Joint.

## Discussion

Prophylaxis is the standard of care in the management of haemophlia and has been linked to superior outcomes which may not be achieved without adherence to treatment.^14,15,16^ Adherence among PWH in Nigeria and in sub-Saharan Africa has not been evidenced so we designed this study to evaluate self-reported adherence among PWH and their determinants. Our study shows a high adherence status of 81% among PWH on prophylaxis. While it corroborates the findings from other researchers who reported similar adherence rates, it is contrary to some others that recorded either higher or lower adherence rates.^17,18,19^The disparities may be due to differences in health care models. Whereas the Spanish health system is that of universal free access to health care which may make them less compliant to treatment protocol,^20^Nigerian patients pay out-of-pocket for most health care services and are more likely to feel the financial burden directly. The treatment protocol used may also be a factor. Dover et al provided evidence to show that PWH with less frequent infusions/week were more likely to adhere to treatment protocol than those who receive more frequent dosing per week.^21^We use a two-dose-per-week dosing for our PWH on low dose prophylaxis. Knowledge also influences the level of adherence to medications.^22^Haemophilia treatment centres whose health care professionals are more knowledgeable about the dosing, dosage and effects of the medications are more likely to have high adherence than those who are not because they are more likely to impart positively to medication compliance by recommending a better protocol and handling drug-related problems.^22^Even though we did not assess knowledge, it could have accounted for the higher adherence obtained by The total mean score for the VERITAS-Pro subscales is 37.35 and 63.0 for the adherence and non-adherence groups respectively. This is comparable to previous studies. Similarly, the subscales “timing”, “skipping”, “planning”, “remembering”, and “communication” where significantly different between groups. Only dosing did not differ suggesting that those in non-adherence group could have skipped infusion, failed to maintain stock at home, had poor communication with their treatment centres, or altered their dosing timing, but maintained the recommended dose of infusion.

Planning, one of the VERITAS-Pro subscales had the least frequency among the PWH in adherence group, only 20 (47.6%) of the adherence group had good planning, the problem could stem from not having enough factor at home and not keeping track of what is left to eventually running out of factors. More efforts should be made to maintain optimal stock of factors at home. For the non-adherent group, the subscale “remember” was the least. This indicates non-intentional non-adherence and it is similar to the work done by van Os et al.^17^ The problem could have been forgetting to infuse the available factor even though they generally had poor planning among other subscales.

Interestingly, we observed that the mean age for those in adherence group was significantly higher than those in non-adherence group which may suggest that within our cohort of PWH, adherence is poorer among the younger age group. This is contrary to the reports by previous studies conducted among PWH.^23^Prophylaxis is usually delivered to the peadiatric/younger age group by caregivers. In our own case, the disparity may be a function of caregiver burden and economic strain which were not assessed.^8^Challenges of the caregiver in balancing the care offered to the child versus other family members as well as social needs could have played out in our study. Other possible reason supporting a lower adherence in younger children in our study could be that of venous access.^24^Securing a venous access in children is more challenging than that of adults or older children and obtaining a central line access is not a common practice among our patients as both caregivers and patients are not favourably disposed to it.

The purpose of instituting prophylaxis in haemophilia management is to prevent symptoms and attendant complications. There is evidence to support that PWH have reduced bleeds, pains, absenteeism and improved quality of life. Our study however did not show any difference in the mean ABR and number of target joints between the adherence and non-adherence groups. This is similar to the report by Krishnan on paediatric patients. All the participants had at least one bleeding episode and a target joint in the preceding year regardless of being on prophylaxis. Despite the lack of association between adherence and ABR, the skipping subscale showed a relationship such that targeting the skipping component may lead to improvements in ABR.

Skipping may suggests that the PWH were intentional about non-adeherence.^25^ For instance, PWH may skip infusion in the absence of bleeds and still score for skipping in the VERITA-Pro study. Only subscale “skipping” showed an association with adherence for ABR. The subscales “timing”, “dosing”, “planning”, “remembering”, and “communication” did not show any association with adherence status for both ABR and number of target joints. This may be due to variations in daily routine.^21^ PWH may make adjustments in their dosing without communicating with the treatment centre, for example, a dose slated to be taken in the morning may be self-rescheduled for evening to accommodate personal program.

The results of this cross-sectional study should be interpreted in the light of its limitations. Being a self-reported adherence study, participants may be faced with recall bias and must have missed to accurately report some clinical outcomes. Our inability to separate the PWH into paediatrics, young adolescents and adults due to small sample size may pose a limitation.

## Conclusion

the findings indicate that adherence was good, and most were in communication with their treatment centers. The skipping subscale was significantly associated with ABR for the nonadherent group. Interventions aimed at improving adherence are the key to better treatment outcomes. A robust multicenter study was needed to assess the reason for poor adherence.

## Data Availability

All relevant data are within the manuscript and its Supporting Information files.

## Acknowledgement

– we acknowledge and thank Natalie A Duncan at the Indiana Haemophilia and Thrombosis Centre for sharing the Validated Haemophilia Regimen-Prophylaxis (VERITAS-Pro).

